# Connecting the Dots: Systematic Exploration of COVID-19 and Acute Kidney Injury through Meta-Analysis

**DOI:** 10.1101/2023.12.01.23299310

**Authors:** Md. Safiullah Sarker, Rubiyat Jahan

## Abstract

**Objective:** COVID-19 pandemic is a danger for the whole world. Also, our knowledge about acute kidney injury (AKI) in COVID-19 patients is incomplete. Few studies informed that the problem of AKI is a common complication, but other studies concluded that AKI is only an unusual event during COVID-19 infection. This study using meta-analysis tools aimed to find disease progression and mortality risk in affected population.

**Methods:** We systematically reviewed the literature on COVID-19 and its association with AKI as per PRISMA guideline. All authors independently performed a literature search until 8th June 2023. We included studies which reported clinical characteristics, incidence of AKI, and the death risk with AKI during COVID-19 infection.

**Findings:** We have included five studies and all of them reported older age (73-75) and males (67-84.2%) were risk factors for patient illness. COVID-19 patients with AKI had more than five times mortality risk of those without AKI. Diagnosis time after disease onset was 8.5 days (IQR, [4–11]). Fatality time after initial hospital admission was 13.5 days (IQR, 8–17). In non-survivors, systemic inflammation with high temperature, abnormal respiratory rate, acute myocardial injury, and acute respiratory distress syndrome (ARDS) were observed. Abnormal biochemical analytes and immunological markers were observed.

**Conclusion:** Our analyses indicate that patients experienced repeated changes in biochemical analytes and immune marker with the progression of the disease. It indicates the requirement of early management and treatment. Further study is required to conclude and to have better knowledge of AKI mechanism with COVID-19 infection.

## Introduction

As per worldometer report as of 8^th^ June, 2023 there were 690,140,273 Coronavirus positive cases and 6,896,133 people were died [1]. The Coronavirus 2019 (COVID-19) disease outbreak occurred in Wuhan, Hubei Province, China, in December 2019, and rapidly spread to other areas of the worldwide. Now, it is well known that the novel Coronavirus is more dangerous than a common respiratory virus. Usually it targets the lungs, but it can attack the whole body. Physician observed that it can cause blood clot and multi organ failure. It was observed that a certain portion of COVID-19 affected patient develops acute kidney injury with normal creatinine level [6].

The SARS-CoV-2 virus affect the respiratory organ but based on clinical findings of several deceased cases the involvement of other organs needs to be explored [6]. Since information on kidney disease in patients with COVID-19 is limited, in this meta-analysis we want to determine the incidence of acute kidney injury (AKI) in patients with COVID-19 by pooling the available published data. Further, we evaluated the association between markers of abnormal kidney function and death in patients with COVID-19. Now, there are still no specific treatments or vaccines for COVID-19 affected patient. We want to discuss possible pathogenic mechanism which may shed light on future intervention.

## Material and methods

We systematically reviewed the literature on COVID-19 and its association with acute kidney injury symptoms. The systematic review was performed according to the Preferred Reporting Items for Systematic Reviews and Meta-Analyses (PRISMA) statement (Table 1)[2].

**Table 1.**
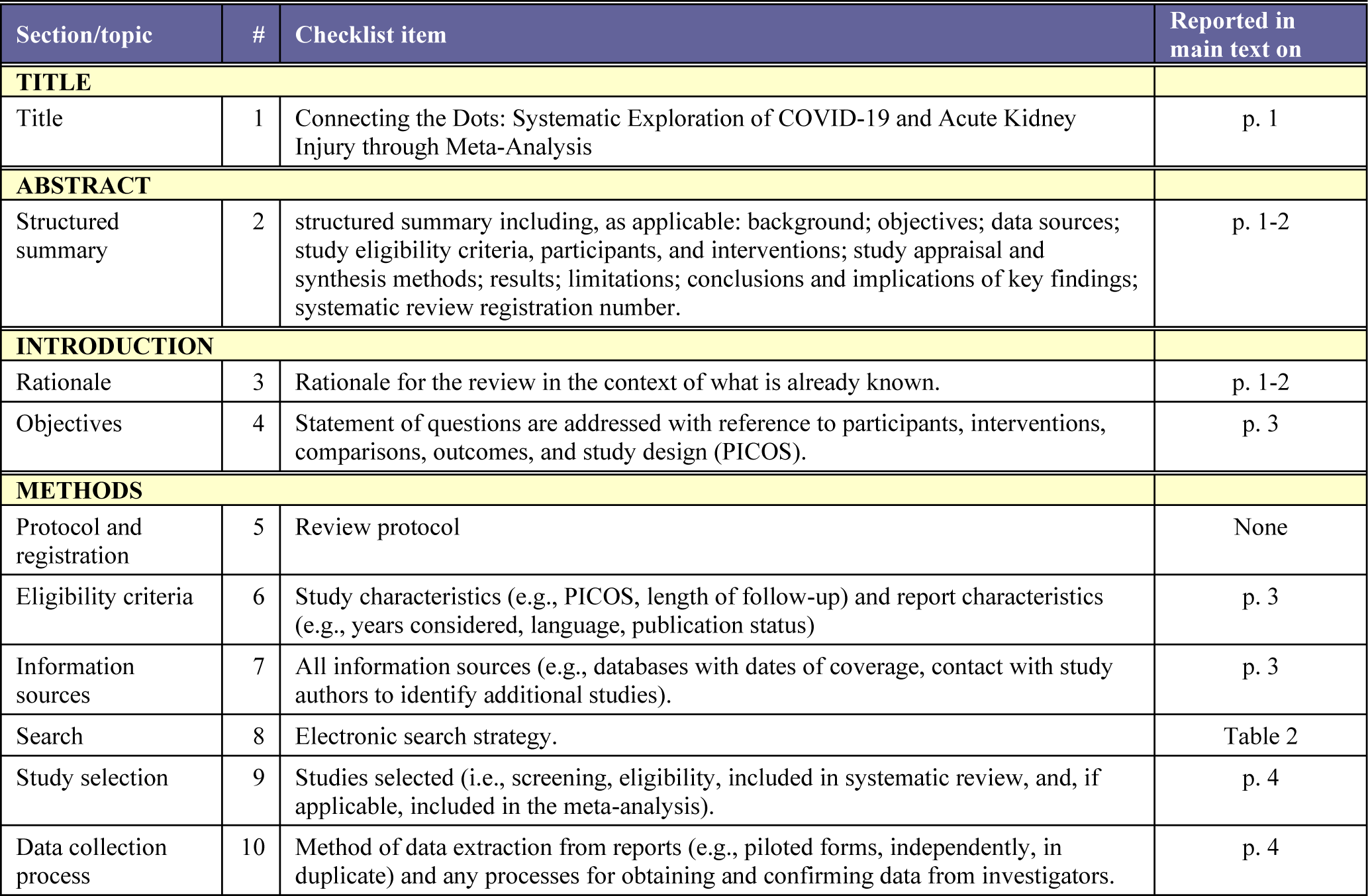
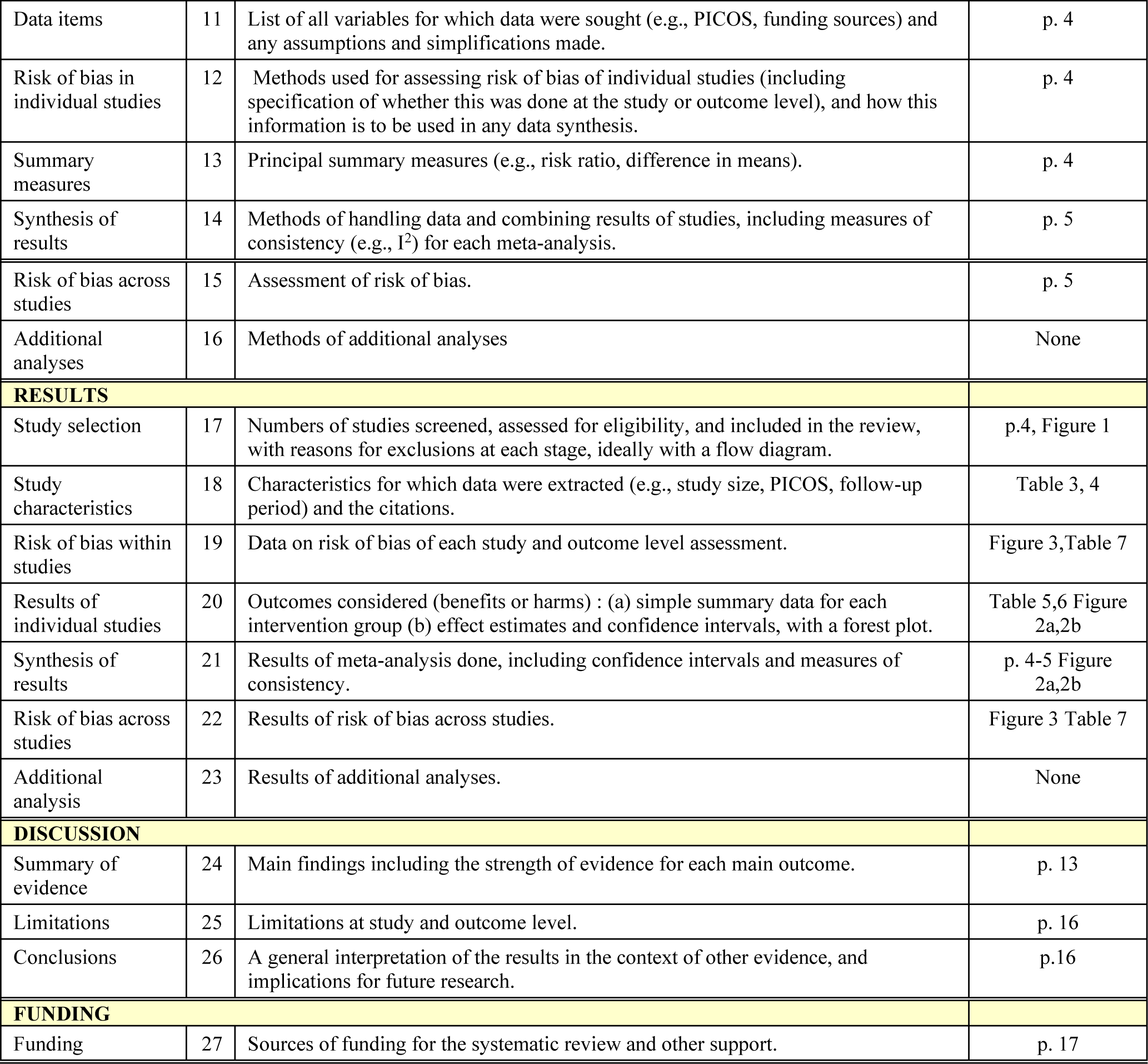
Preferred Reporting Items for Systematic Reviews and Meta-analyses (PRISMA) checklist.

Two authors independently performed a literature search using PubMed, Web of Science, Embase, and Cochrane Library fulfilled the pre-specified criteria until 8th June, 2023 to include studies reported the necessary clinic characteristics, the incidence of AKI, the mortality with AKI and the death risk with AKI during COVID-19 infection were pooled for statistical analysis by MedCalc software [3] to get conclusions.

### Literature search

A comprehensive literature search was performed using combination of keywords (MeSH terms and free text words) including ‘COVID-19’/ ‘SARS-CoV-2’, ‘AKI’/‘acute kidney injury’, and “English language”. PubMed, Embase, Cochrane library, Cochrane COVID-19 Study Register and DIRECTORY OF OPEN ACCESS JOURNALS(DOAJ) and Web of Science were searched up to 8th June 2023. The search strategy was present in Table 2. Additional articles were sought from the reference lists of the included studies.

**Table 2.** Search criteria for systematically reviewing COVID-19 and Acute kidney injury (AKI) in PubMed, Embase Cochrane Library, Cochrane COVID-19 Study Register and Web of Science.

| ID | Search site | Key word | Search terms | Hits |
| --- | --- | --- | --- | --- |
| #1 | PubMed | COVID-19 | (((((("covid 19"[All Fields] OR "covid 2019"[All Fields]) OR "severe acute respiratory syndrome coronavirus 2"[Supplementary Concept]) OR "severe acute respiratory | 17,850 |
| #2 | PubMed | (COVID-19) AND (Acute Kidney Injury) | (((((("covid 19"[All Fields] OR "covid 2019"[All Fields]) OR "severe acute respiratory syndrome coronavirus 2"[Supplementary Concept]) OR "severe acute respiratory syndrome coronavirus 2"[All Fields]) OR "2019 ncov"[All Fields]) OR "sars cov 2"[All Fields]) OR "2019ncov"[All Fields]) OR ((("wuhan"[All Fields] AND ("coronavirus"[MeSH Terms] OR "coronavirus"[All Fields])) AND (2019/12/1:2023/05/31[Date - Publication] OR 2020/1/1:2023/05/31[Date - Publication]))) AND ((("acute kidney injury"[MeSH Terms] OR ("acute"[All Fields] AND "kidney"[All Fields]) AND "injury"[All Fields])) OR "acute kidney injury"[All Fields]) | 78 |
| #3 | PubMed | Acute Kidney Injury) AND (SERS-CoV-2) | ("acute kidney injury"[MeSH Terms] OR ((("acute"[All Fields] AND "kidney"[All Fields]) AND "injury"[All Fields])) OR "acute kidney injury"[All Fields] | 62063 |
| #4 | PubMed | ((Acute Kidney Injury) AND (SERS-CoV-2)) OR (COVID-19) | ((("acute kidney injury"[MeSH Terms] OR ((("acute"[All Fields] AND "kidney"[All Fields]) AND "injury"[All Fields])) OR "acute kidney injury"[All Fields]) OR ((((((("covid 19"[All Fields] OR "covid 2019"[All Fields]) OR "severe acute respiratory syndrome coronavirus 2"[Supplementary Concept]) OR "severe acute respiratory syndrome coronavirus 2"[All Fields]) OR "2019 ncov"[All Fields]) OR "sars cov 2"[All Fields]) OR "2019ncov"[All Fields]) OR ((("wuhan"[All Fields] AND ("coronavirus"[MeSH Terms] OR "coronavirus"[All Fields])) AND (2019/12/1:2023/05/31[Date - Publication] OR 2020/1/1:2023/05/31[Date - Publication])))) | 79835 |
| #5 | PubMed | Acute Kidney Injury | ("acute kidney injury"[MeSH Terms] OR ((("acute"[All Fields] AND "kidney"[All Fields]) AND "injury"[All Fields])) OR "acute kidney injury"[All Fields] | 62063 |
| #6 | PubMed | SERS-CoV-2 | it retrieved zero results. | 0 |
| #7 | PubMed | AKI | ("AKI"[All Fields] | 13853 |
| #8 | PubMed | (Coronavirus disease 2019) AND (acute kidney injury) | ((("covid 19"[Supplementary Concept] OR "covid 19"[All Fields]) OR "coronavirus disease 2019"[All Fields]) AND ((("acute kidney injury"[MeSH Terms] OR ((("acute"[All Fields] AND "kidney"[All Fields]) AND "injury"[All Fields])) OR "acute kidney injury"[All Fields]) | 85 |
| #9 | PubMed | ((Coronavirus disease 2019) AND (acute kidney injury)) AND (English[Language]) | ((("covid 19"[Supplementary Concept] OR "covid 19"[All Fields]) OR "coronavirus disease 2019"[All Fields]) AND (("acute kidney injury"[MeSH Terms] OR ("acute"[All Fields] AND "kidney"[All Fields]) AND "injury"[All Fields])) OR "acute kidney injury"[All Fields]) AND "English"[Language]) | 81 |
| #10 | PubMed | ((COVID-19) AND (acute kidney injury)) AND (English[Language]) | ((((((("covid 19"[All Fields] OR "covid 2019"[All Fields]) OR "severe acute respiratory syndrome coronavirus 2"[Supplementary Concept]) OR "severe acute respiratory syndrome coronavirus 2"[All Fields]) OR "2019 ncov"[All Fields]) OR "sars cov 2"[All Fields]) OR "2019ncov"[All Fields]) OR (("wuhan"[All Fields] AND ("coronavirus"[MeSH Terms] OR "coronavirus"[All Fields])) AND (2019/12/1:2023/05/31[Date - Publication] OR 2020/1/1:2023/05/31[Date - Publication]))) AND (("acute kidney injury"[MeSH Terms] OR ("acute"[All Fields] AND "kidney"[All Fields]) AND "injury"[All Fields])) OR "acute kidney injury"[All Fields]) AND "English"[Language]) | 82 |
| #11 | PubMed | ((novel coronavirus disease) AND (acute kidney injury)) AND (English[Language]) | ((("novel"[All Fields] OR "novel s"[All Fields]) OR "novels"[All Fields]) AND (("coronavirus"[MeSH Terms] OR "coronavirus"[All Fields]) OR "coronaviruses"[All Fields]) AND (((("disease"[MeSH Terms] OR "disease"[All Fields]) OR "diseases"[All Fields]) OR "disease s"[All Fields]) OR "diseased"[All Fields])) AND (("acute kidney injury"[MeSH Terms] OR ("acute"[All Fields] AND "kidney"[All Fields]) AND "injury"[All Fields])) OR "acute kidney injury"[All Fields]) AND "English"[Language]) | 24 |
| #12 | PubMed | ((COVID-19) AND (acute kidney injury)) AND (English[Language]) OR ((novel coronavirus disease) AND (acute kidney injury)) AND (English[Language]) | ((((((("covid 19"[All Fields] OR "covid 2019"[All Fields]) OR "severe acute respiratory syndrome coronavirus 2"[Supplementary Concept]) OR "severe acute respiratory syndrome coronavirus 2"[All Fields]) OR "2019 ncov"[All Fields]) OR "sars cov 2"[All Fields]) OR "2019ncov"[All Fields]) OR (("wuhan"[All Fields] AND ("coronavirus"[MeSH Terms] OR "coronavirus"[All Fields])) AND (2019/12/1:2023/05/31[Date - Publication] OR 2020/1/1:2023/05/31[Date - Publication]))) AND (("acute kidney injury"[MeSH Terms] OR ("acute"[All Fields] AND "kidney"[All Fields]) AND "injury"[All Fields])) OR "acute kidney injury"[All Fields]) AND "English"[Language]) OR (((("novel"[All Fields] OR "novel s"[All Fields]) OR "novels"[All Fields]) AND (("coronavirus"[MeSH Terms] OR "coronavirus"[All Fields]) OR "coronaviruses"[All Fields]) AND (((("disease"[MeSH Terms] OR "disease"[All Fields]) OR "diseases"[All Fields]) OR "disease s"[All Fields]) OR "diseased"[All Fields])) AND (("acute kidney injury"[MeSH Terms] OR ("acute"[All Fields] AND "kidney"[All Fields]) AND "injury"[All Fields])) OR "acute kidney injury"[All Fields]) AND "English"[Language]) | 88 |
| #13 | PubMed | ((((COVID-19) AND (acute kidney injury)) AND (English[Language])) OR ((novel coronavirus disease) AND (acute kidney injury)) AND (English[Language])) | ((((((((((("covid 19"[All Fields] OR "covid 2019"[All Fields]) OR "severe acute respiratory syndrome coronavirus 2"[Supplementary Concept]) OR "severe acute respiratory syndrome coronavirus 2"[All Fields]) OR "2019 ncov"[All Fields]) OR "sars cov 2"[All Fields]) OR "2019ncov"[All Fields]) OR (("wuhan"[All Fields] AND ("coronavirus"[MeSH Terms] OR "coronavirus"[All Fields])) AND (2019/12/1:2023/5/31[Date - Publication] OR 2020/1/1:2023/05/31[Date - Publication])))) AND ((("acute kidney injury"[MeSH Terms] OR ("acute"[All Fields] AND "kidney"[All Fields]) AND "injury"[All Fields]) OR "acute kidney injury"[All Fields]) AND "English"[Language]) OR (((("novel"[All Fields] OR "novel s"[All Fields]) OR "novels"[All Fields]) AND ("coronavirus"[MeSH Terms] OR "coronavirus"[All Fields] OR "coronaviruses"[All Fields]) AND (((("disease"[MeSH Terms] OR "disease"[All Fields]) OR "diseases"[All Fields]) OR "disease s"[All Fields]) OR "diseased"[All Fields])) AND ((("acute kidney injury"[MeSH Terms] OR ("acute"[All Fields] AND "kidney"[All Fields]) AND "injury"[All Fields]) OR "acute kidney injury"[All Fields]) AND "English"[Language])) | 84 |
| #14 | Cochrane Library | COVID-19 AND AKI | (COVID-19):ti,ab,kw AND (Acute Kidney Injury):ti,ab,kw | 5 |
| #15 | Cochrane Library | COVID-19 AND Acute Kidney Injury | (COVID-19 AND Acute Kidney Injury):ti,ab,kw (Word variations have been searched) | 6 |
| #16 | Cochrane COVID-19 Study Register | COVID-19 and acute kidney injury | COVID-19 and acute kidney injury | 1,711 |
| #17 | DIRECTORY OF OPEN ACCESS JOURNALS(DOAJ) | COVID-19 and acute kidney injury | COVID-19 and acute kidney injury | 13 |
| #20 | Embase | COVID-19 AND acute kidney injury | "COVID-19 AND acute kidney injury" in journals | 2,360 |
| #21 | Web of Science | COVID-19 AND acute kidney injury | (from Web of Science Core Collection) | 24 |

### Selection criteria

All articles identified from the literature search were screened by two independent reviewers (M.S.S and R.J). We included studies on COVID-19 in adult patients. Full articles, Conferences abstracts, Case reports, case series, non-randomized studies, randomized trials and observational studies, that were published in English were included in this review. Commentaries, letters to editors and editorials were also included. Studies on severe acute respiratory syndrome (SARS) and Middles East respiratory syndrome (MERS) but not COVID-19 were excluded.

### Data Collection

For eligible studies, study information including first authors, site of study, inclusion and exclusion criteria, sample size, age and sex were recorded. A standardized form for data entry has been devised to focus on the following areas: number of patients, number of AKI patients, AKI related indicators (according to Acute Kidney Injury Network (AKIN): (a) an increase in the serum creatinine level to ≥0.3 mg/dL, (b) an increase in baseline serum creatinine level to ≥150%)[4], AKI related death (Table 3 and 4).

**Table 3.** Characteristics of studies included in the systematic review (qualitative analysis).

| SL# | Study name | Reference# | Journal | Date of Publication | Study Population (n) | Age in Years (range) | Sex |  |
| --- | --- | --- | --- | --- | --- | --- | --- | --- |
|  |  |  |  |  |  |  | M, n (%) | F, n (%) |
| 1 | Lim et al., | 4 | J. Clin. Med | 3 June 2020 | 160 | 69.0 (24.0-98.0) | 88 (55) | 72 (55) |
| 2 | Shao et al., | 5 | Front. Med | 22 May 2020 | 155 | 48.0 (7.0–96.0) | 62(40) | 93(60) |
| 3 | Cheng et al., | 6 | Kidney International | 16 March 2020 | 701 | 63.0 (50.0–71.0) | 367(52.4) | 334(47.7) |
| 4 | Wang et al | 7 | Critical Care | 30 April 2020 | 107 | 51.0 (19.0–92.0) | 57 (53.3) | 44(46.7) |

**Table 4:** Characteristics of studies included in the systematic review (Quantitative analysis)

| Reference | Authors | Journal | Date of Publication | Study population | AKI definition | AKI related death | AKI | Baseline creatinine | Blood Urea Nitrogen (BUN) | Proteinuria | Hematuria | eGFR | D-dimer | RRT(n) |
| --- | --- | --- | --- | --- | --- | --- | --- | --- | --- | --- | --- | --- | --- | --- |
| 4 | Lim et al., | J. Clin. Med | 3 June 2020 | 164 | AKIN | 17 | 30 | 30/30 | 30/160 | N/A | N/A | 30/30 | 16/30 | CRRT (5) |
| 5 | Shao et al., | Front. Med | 22 May 2020 | 155 | N/A | 7 | 7 | 18/155 | 18/155 | N/A | N/A | N/A | 18/155 | CRRT (7) |
| 6 | Cheng et al., | Kidney International | 16 March 2020 | 701 | KDIGO | HR | 36 | 36/36 | 701 | 194/442 | 118/442 | 701/701 | 512/661 | N/A |
| 7 | Wang et al | Critical Care | 30 April 2020 | 107 | KDIGO | OR | 14 | 107 | N/A | N/A | N/A | N/A | 107 | CRRT (4) |
| 8 | Li et al. | The Lancet Infectious Diseases | 2020 | 193 | KDIGO | HR | 55 | 43/193 | 59/193 | 88/147 | 71/147 | N/A | 128/182 | CRRT (7) |

Abbreviations: AKI, acute kidney injury; AKIN, Acute Kidney Injury Network; RRT, Renal Replacement Therapy, (n) indicate number of patients received RRT, CRRT, continuous renal replacement therapy; KDIGO, Kidney Disease: Improving Global Outcomes; HR, Hazard Ratio, OR, Odd Ratio,

### Data synthesis and statistical analysis

Primary outcomes of our study included acute kidney injury (AKI) in COVID-19 infected patient (Table 5), death of AKI developed patients (Table 6), clinical symptoms, markers of kidney injury. For the AKI prevalence rates, data were pool analyzed using SPSS and MedCalc. When there are two or more studies with at least four patients reporting the same outcome under the same definition, random effects model was used. Double arcsine transformation was used to pool the proportions. The results were presented by forest plots (Figure 2a,b), illustrating the prevalence, 95% confidence intervals and weightings. Cochran’s Q test was used to detect heterogeneity, and a *p* value of < 0.10 indicates significant heterogeneity. ^I2^ statistics was calculated to measure the proportion of total variation in study estimates attributed to heterogeneity with I^2^> 50% suggests substantial heterogeneity.

**Table 5.** Incidence of AKI patient those who were SARS-CoV-2 positive.

| SL # | Study name | Ref# | AKI(n) | Total Death (n) | Non-AKI(n) | Death in non-AKI (n) | Total | Prevalence (%) | Weight (%) | Risk Ratio |
| --- | --- | --- | --- | --- | --- | --- | --- | --- | --- | --- |
| 1 | Lim et al.,2020 | 4 | 17 | 30 | 134 | 13 | 164 | 10.37 | 12.42 | 6.6 |
| 2 | Shao et al.,2020 | 5 | 7 | 18 | 137 | 11 | 155 | 4.52 | 11.74 | 5.1 |
| 3 | Cheng et al., 2020 | 6 | 36 | 113 | 588 | 77 | 701 | 13.08 | 8.11 | 15.0 |
| 4 | Wang et al, 2020 | 7 | 14 | 19 | 88 | 5 | 107 | 5.14 | 53.11 | 2.6 |
| 5 | Li et al.,2020 | 8 | 32 | 55 | 138 | 23 | 170 | 28.50 | 14.62 | 4.3 |
|  | Total |  | 106 | 235 | 1085 | 129 | 1320 | 8 | 100 | 4.1 |

**Table 6.** Death in acute kidney injury patient who were SARS-COV-2 infected.

| # | Study name | Ref# | Death (n) | AKI(n) | Prevalence (%) | Weight (%) |
| --- | --- | --- | --- | --- | --- | --- |
| 1 | Lim et al., | 4 | 17 | 30 | 56.67 | 18.99 |
| 2 | Liang et al., | 5 | 7 | 18 | 38.89 | 11.39 |
| 3 | Cheng et al., | 6 | 6 | 36 | 16.67 | 22.78 |
| 4 | Wang et al., | 7 | 14 | 19 | 73.68 | 12.03 |
| 5 | Li et al. | 8 | 32 | 55 | 58.18 | 34.81 |

**Figure 2a:**
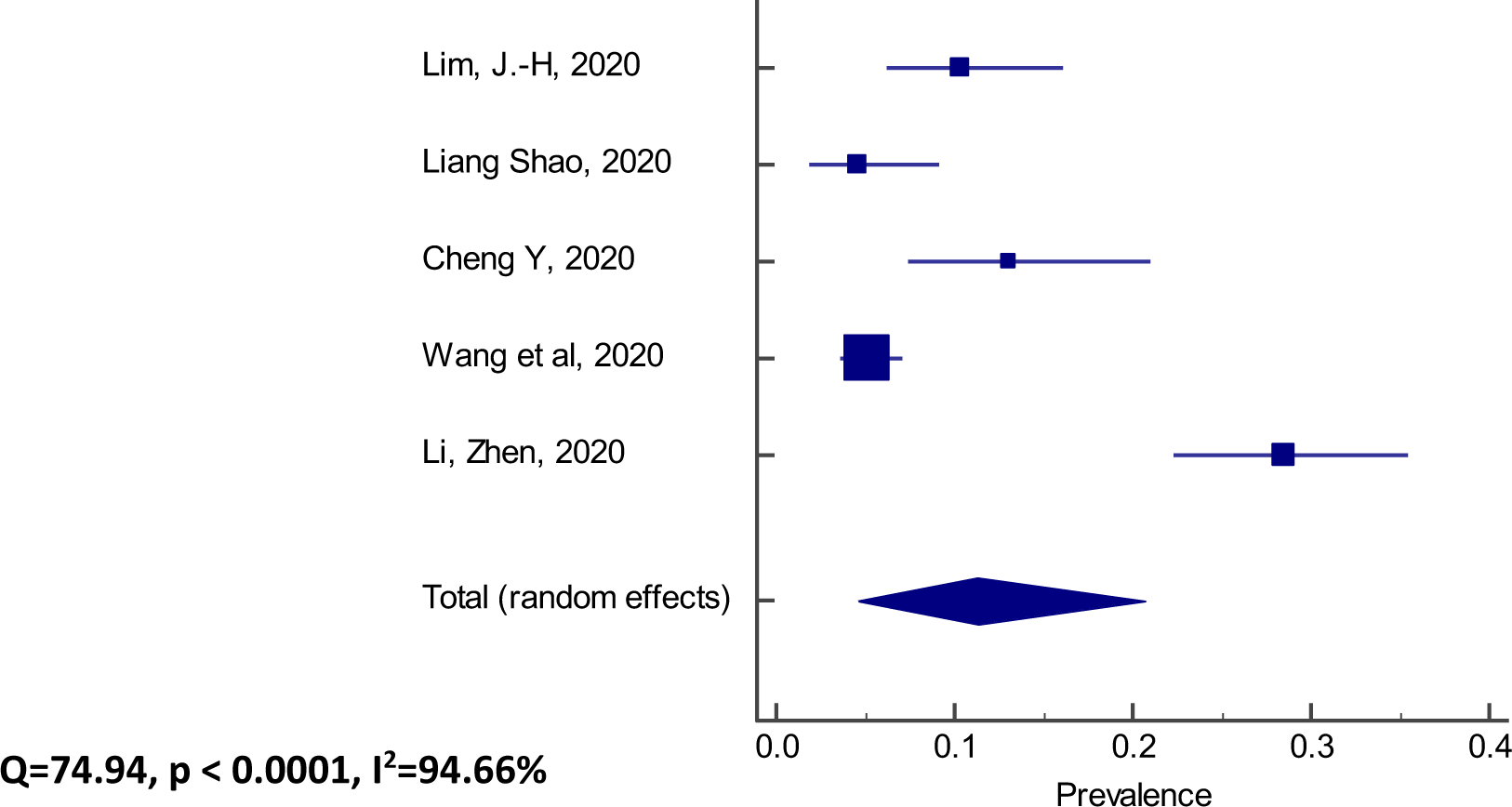
Prevalence of Death in AKI patient those who were SERS-CoV-2 positive

**Figure 2b:**
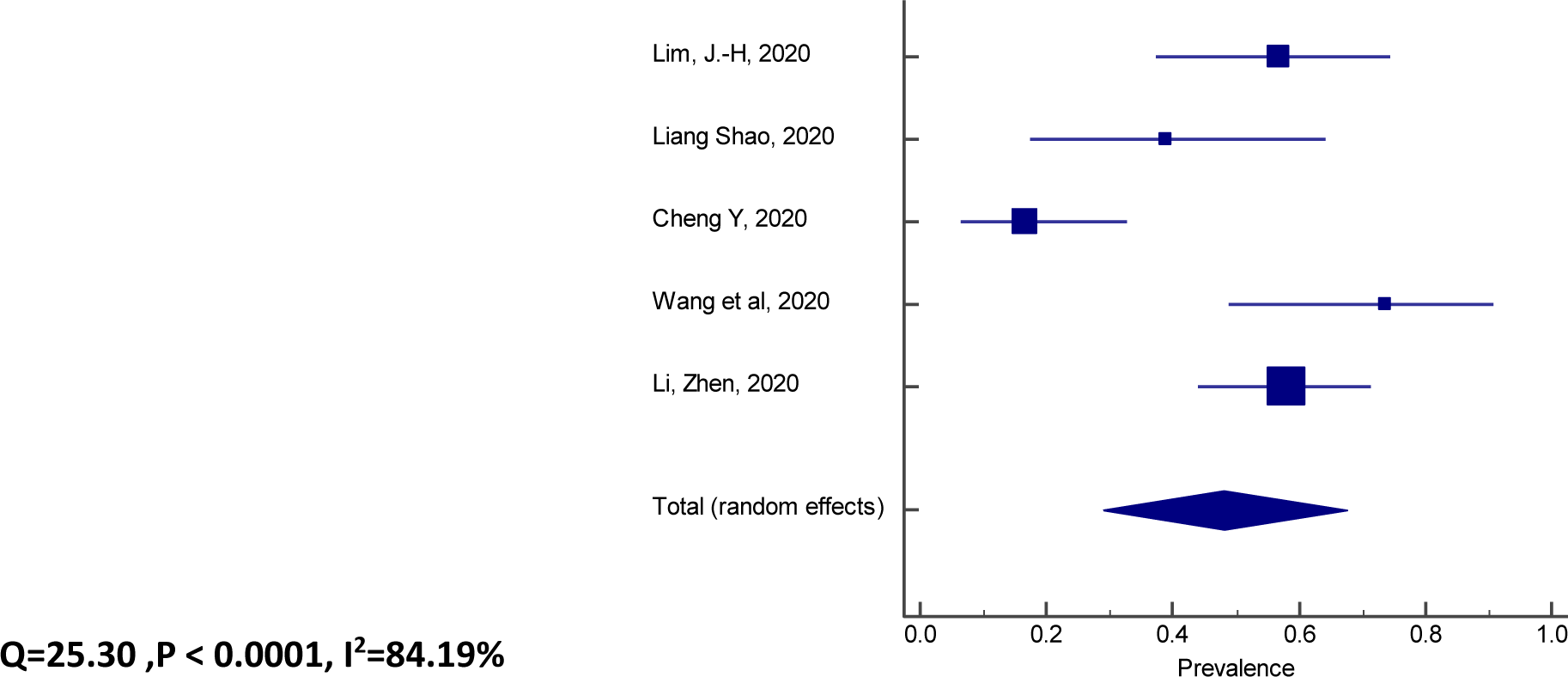
Death in acute kidney injury patient who were SERS-COV-2 infected

## Results

The PRISMA flow diagram was shown in Figure 1. A total of 1723 records were identified upon literature search, and 11 additional records were sought from reference lists of the included literature. 1535 records remained after removing duplicates. Among them, 757 were screened and 816 were excluded (641 not related and 135 review articles). Of the screened articles 108 were assessed for eligibility (COVID-19 and AKI) and 649 were excluded as not related to AKI. Of the 108 articles only 5 were eligible for serum creatinine marker. These five articles were selected for quantitative analysis [4–8]. Using funnel plot one study was excluded and the remaining four were selected for qualitative analysis. Table 3 give a clear picture of basic characteristics of the 4 studies [4,6–8] that were included in the qualitative analysis. The Newcastle-Ottawa score of the included studies is shown in Table 7.

Results of quality assessment for risk of selection bias as per Newcastle-Ottawa Scale.

**Table 7. Results of quality assessment for risk of selection bias as per Newcastle-Ottawa Scale**
| Author, year | Age | Gender | Sampling method | Location | Author, year | Total scores | Risk of selection bias |
| --- | --- | --- | --- | --- | --- | --- | --- |
| Lim, J.-H,2020 | 1 | 1 | 1 | 1 | 1 | 5 | Low risk |
| Liang Shao,2020 | 1 | 1 | 1 | 1 | 1 | 5 | Low risk |
| Cheng Y,2020 | 1 | 1 | 1 | 1 | 1 | 5 | Low risk |
| Wang et al,2020 | 1 | 1 | 1 | 1 | 1 | 5 | Low risk |

Publication bias was shown in the funnel plot analysis in Figure 3.

**Figure 3.**
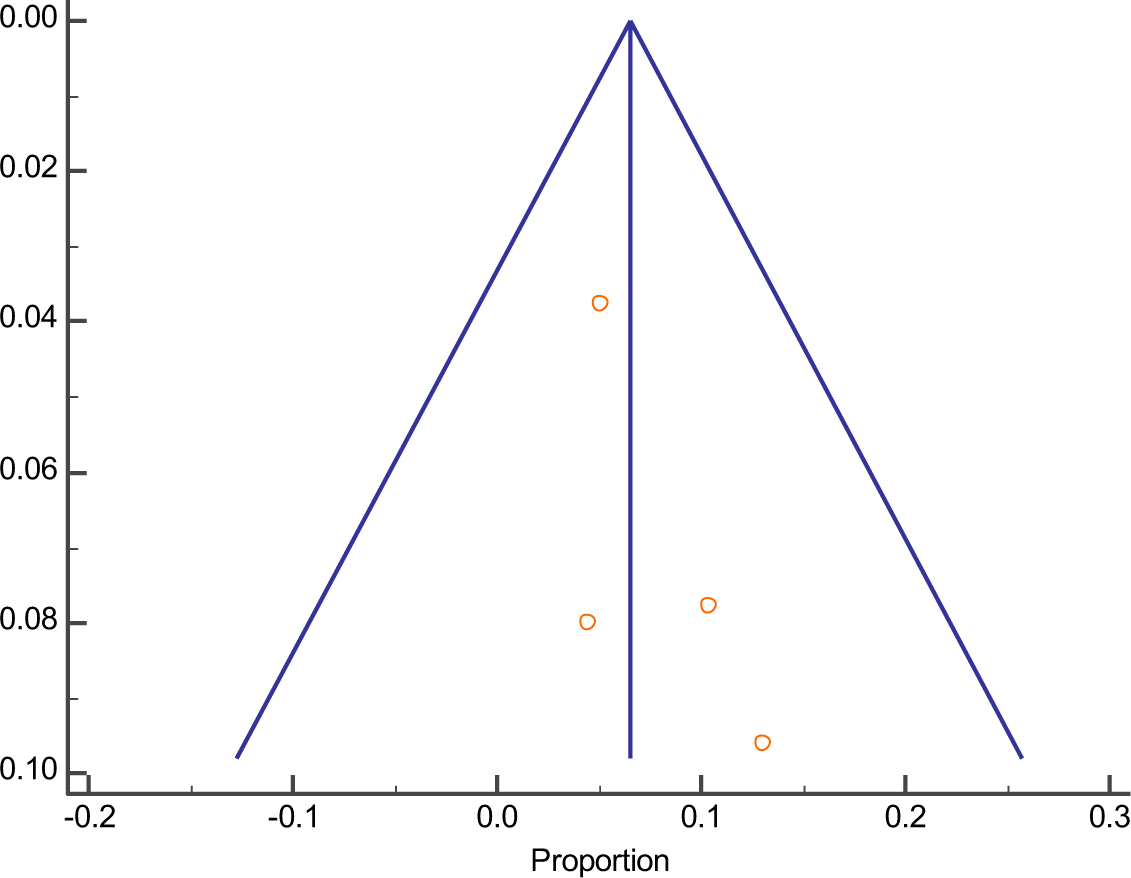
Funnel plot to select study for qualitative analysis

There was no evidence of publication bias when applying funnel plot analysis (Figure 3). As per our meta-analysis it was observed that mortality is remarkably higher in COVID-19 infected patients with severe AKI. At present, scientists observed the incidence of AKI among patients with COVID-19 with varying conclusion. It was observed to occur nearly one in four of patients with COVID-19 [9]. Our meta-analysis included five studies and the findings are discussed below:

### High mortality in older male patient

We have included five studies for our analysis. Of these five all of them reported older age and male sex were risk factors for patient illness. We were able to extract data from three studies. The study by Lim et al., reported of the non-survivor median age was 75.0 years and about 67% were male [4]. Shao et al., observed that of the non-survivors, the median age was 73.5 years, and the proportion of males was 72.2% [5]. Wang et al., also reported the same findings [of the non-survivors(n=19) were of 73.0 years and 84.2% (16/19) were male] [7].

### In-hospital mortality was higher in AKI patients

All of the studies observed most case fatality in AKI patient specially in stage III AKI group who needed renal replacement therapy. Study of Li et al. reported by Cox regression analysis that COVID-19 patients who developed AKI had a ∼5.3-times mortality risk of those without AKI [8].

### The interval between disease onset and Diagnosis

The incubation period of COVID-19 is considered to be up to14 days following exposure [10–12]. Two studies highlighted on this issue. Cheng et al and Wang et al., define the date of disease onset was the day when the first symptom was noticed [7]. Wang et al also informed that “median duration from illness onset to admission was 10 days”. Shao et al.; observed that ‘diagnosis time after disease onset was 8.5 days (IQR, [4–11]) [5]. Lim et al. reported that there were 17 deaths among AKI patients; [stage-I: 4 (28.6%), stageII: 1 (25.0%), and stageIII: 12 (100.0%), respectively] [4]. It was also mentioned in the same article that in-hospital mortality was higher in AKI patients than in non-AKI patients (56.7% vs. 20.8%, p < 0.001) [ 4]. It was observed that there was more death in stage 3 AKI than non-AKI or stage 1 AKI (hazard ratio (HR) = 3.62 (95% confidence interval (CI) = 1.75–7.48), p = 0.001; HR = 15.65 (95% CI = 2.43–100.64), p = 0.004). Using Kaplan–Meier analysis Cheng et.al, showed that patients with kidney disease had a significantly higher risk for in-hospital death. Cox proportional hazard regression established that AKI stage 1(hazard ratio: 1.90, 95% confidence interval: 0.76-4.76), stage 2 (3.51, 1.49-8.26), stage 3 (4.38, 2.31-8.31) [6].

### Fatality time

Two studies reported about the fatality time form onset of illness to death. Shao et al observed a “period of 7–13 days after illness onset is the critical stage in the COVID-19 course. All patients died within 45 days after the initial hospital admission with a median survivor time of 13.5 days (IQR, 8–17)” [5]. Cheng et al., also support similar findings and observed that the prevalence of kidney disease on admission and the development of AKI during hospitalization in patients with COVID-19 is high and is associated with in-hospital mortality [6].

### Inflammation in AKI and acute respiratory distress syndrome

Wang et al; observed in non-survivors, the appearance of systemic inflammation with high temperature, abnormal respiratory rate, WBC counts and neutrophil counts. Subsequently, multiple organ dysfunction syndromes (MODS) occurred with thrombocytopenia, renal failure, acute myocardial injury, and ARDS [7].

### Biochemical parameters

The following abnormal biochemical parameter were observed in COVID-19 patient with AKI:

### Elevated baseline serum creatinine

The baseline serum creatinine level was defined as the serum creatinine value on admission or within 6 months before admission [4]. The typical reference range for serum creatinine is 60 to 110 micromoles per liter (μmol/L) (0.7 to 1.2 milligrams per deciliter (mg/dL)) for men and 45 to 90 μmol/L (0.5 to 1.0 mg/dL) for women [13]. Four studies observed elevated serum creatinine in AKI patient [5–8]. Shao et al., observed an elevated serum creatinine level at day of admission and day of death. [88.25 μmol/L (76.70–123.70) and 123.60 (70.80–328.70) respectively, median and IQR]. Cheng Y also observed nearly fifteen percent of the patient on admission had elevated serum creatinine (77±31, mean ± SD). In this study cox proportional hazard regression confirmed that elevated baseline serum creatinine (hazard ratio: 2.10, 95% confidence interval: 1.36-3.26) was independent risk factor for in-hospital death. Wang et al., and Li et al., also observed the same situation.

### Elevated baseline blood urea nitrogen

Blood urea nitrogen (BUN) normal reference range for adults is 3.6-7.1 mmol/L[14]. Cheng et al., observed 13.1% patient has elevated BUN during admission and the whole course of treatment. Cox proportional hazard regression confirmed that elevated baseline blood urea nitrogen (hazard ratio: 3.97, 95% confidence interval: 2.57-6.14), AKI stage 1 (1.90, 0.76-4.76), stage 2 (3.51, 1.49-8.26), stage 3 (4.38, 2.31-8.31) was independent risk factor for in-hospital death. Wang et al., and Li et al., also observed the same situation. Wang et al., observed admission values of blood urea nitrogen were elevated in the non-survivors. Li et al., observed “On hospital admission, a remarkable fraction of patients had signs of kidney dysfunctions, including 14% with increased levels of blood urea nitrogen” [8].

### Estimated Glomerular Filtration Rate (eGFR)

A normal eGFR for adults is greater than 90 mL/min/1.73m2, according to the National Kidney Foundation and actual numbers are only reported once values are less than 60 mL/min/1.73m^2^.[15] The study by Cheng et al., reported during admission 13.1% patients had reduced eGFR (< 60 ml/min/1.73m^2^).

### High level of D-dimer

D-dimer is a protein fragments formed when a blood clot gets dissolved in blood stream. Acute kidney injury (AKI) affects the hypercoagulable state [16] of the body. As coagulation biomarkers D-dimer is measured. It’s normal value is 0–500 mg/L and Shao et al., observed an extremely high level of D-dimer on the death date compared with the admission date (median, 3542.50 [IQR, 2797.00–10929.00] vs. 492.50 [IQR, 273.00–2139.00], p<0.05). Cheng et al., observed an elevated D-dimer on 77.5 % COVID-19 patients during admission. Wang et al., observed the non-survivors had higher D-dimer level (439 mg/L [202–1991]vs 191 mg/L [108–327], P = 0.003) in COVID-19 patients. Li et al., showed that using univariate cox regression analysis, D-dimer were significantly associated with the death of COVID-19 patient [5–8].

### Proteinuria

Patient with proteinuria have urine containing an unusual amount of protein. The condition is a signal of kidney disease. During acute kidney injury albumin leak form the damage kidney filters into the urine [17]. The normal mean albumin excretion rate (AER) is 5-10 mg/day, with an AER of >30 mg/day considered abnormal [18].In this review we included five studies and two of them highlighted on proteinuria in AKI patient. Cheng et al., observed that during hospital admission, almost half of patients had proteinuria. One in twenty patient developed AKI during the study period. Kaplan–Meier analysis confirmed that patients with kidney disease had a top risk for in-hospital death. Cox proportional hazard regression support that proteinuria 1+ (hazard ratio: 1.80, 95% confidence interval: 0.81-4.00), 2+∼3+(4.84, 2.00-11.70), had independent risk factors for in-hospital death [6]. Study by Li et al., also observed during hospital admission, more than half of patients had proteinuria. The univariate cox regression analysis confirms that proteinuria was remarkably associated with the fatality of COVID-19 patients [8].

### Hematuria

Hematuria is defined as the presence of 5 or more red blood cells (RBCs) per high-power field in 3 of 3 consecutive centrifuged specimens available at least 1 week apart. It can be either visible or microscopic. It may also be either symptomatic or asymptomatic, either temporary or constant, and either alone or coupled with proteinuria and other urinary problem [19]. Two of our analyzed studies informed on hematuria in AKI patient. Cheng et al., observed that almost one in four patients had hematuria during hospital admission. During the study period, AKI occurred in almost five percent patients. It was observed that the AKI patient had a top risk for in-hospital mortality by Kaplan–Meier analysis. Cox proportional hazard regression analysis confirmed that hematuria 1+(hazard ratio: 2.99, 95% confidence interval: 1.39-6.42), 2+∼3+ (5.56,2.58-12.01) were independent risk factors for in-hospital death [6]”. Study by Li et al., also support such observation. They showed during hospital admission, nearly half of patients had hematuria. The univariate cox regression analysis confirms that hematuria was remarkably associated with the casualty of COVID-19 patients [8].

### Immune parameters Hematological parameter

Shao et al., observed that “with the deterioration of disease, most patients experienced consecutive changes in lymphopenia, leukocytosis, thrombocytopenia. Regarding the immune dysregulation, patients exhibited significantly decreased T lymphocytes in the peripheral blood, including CD3+T, CD3+CD4+Th, andCD3+CD8+Tc cells”. Wang at el., reported “in severe cases, leukocytosis, neutrophilia, and deteriorating multi-organ dysfunction were dominant” [5]. The role of cytokine is well known by scientific community in viral infection. None of our included studies analyzed cytokine profile. We reviewed literature about cytokine in SARS-CoV-2 infection which is discussed below:

### Cytokine Storm in SARS-CoV-2 Infection

Abnormalities of the inflammatory cytokines profile are good trait during SARS-CoV infection. They correlate with illness but poor prognostic marker [20–21].Many studies observed a group of patients with COVID-19 had inflammatory response and cytokine storm syndrome [22–24]. A cytokine profile with hyper-inflammatory condition defined by a huge cytokine production and serious problem with vital organs failure was observed in COVID-19 patients. In addition, increased ferritin and IL-6 levels observed in more than hundred confirmed COVID-19 cases. It indicates that virus related hyper-inflammation might be one leading cause of serious health problem [25].

In a cohort of 53 patients with COVID-19 compared to healthy controls a marked increase of 14 pro- and anti-inflammatory cytokines were observed. Among them, interferon gamma induced protein 10(IP-10), monocyte chemotactic protein-3(MCP-3) and interleukin (IL-1ra) were significantly associated with disease severity [23]. It supports that abnormal inflammatory cytokine release was dangerous for lung tissue injury and COVID-19 pathogenesis.

### Possible Pathophysiology of AKI in COVID-19

The cause of kidney injury in COVID-19 is thought to be complex [43]. It is related to cardiovascular comorbidity and influencing factors like nephrotoxin, bacteremia and hypovolemia. The right ventricular failure due to COVID-19 pneumonia cause kidney congestion following AKI. Also, left ventricular abnormalities generate low cardiac function, arterial under filling, and kidney hypoperfusion. The endothelium of lung and kidney is disrupted which cause proteinuria. Additionally, virus particles were present in renal endothelial cells as indicated by postmortem report. So, viremia is a possible cause of endothelial damage in the kidney and a probable contributor to AKI [44]. A schematic diagram is drawn in figure 4 to represent the possible pathophysiology of AKI in SARS-CoV-2 infection.

**Figure 4:**
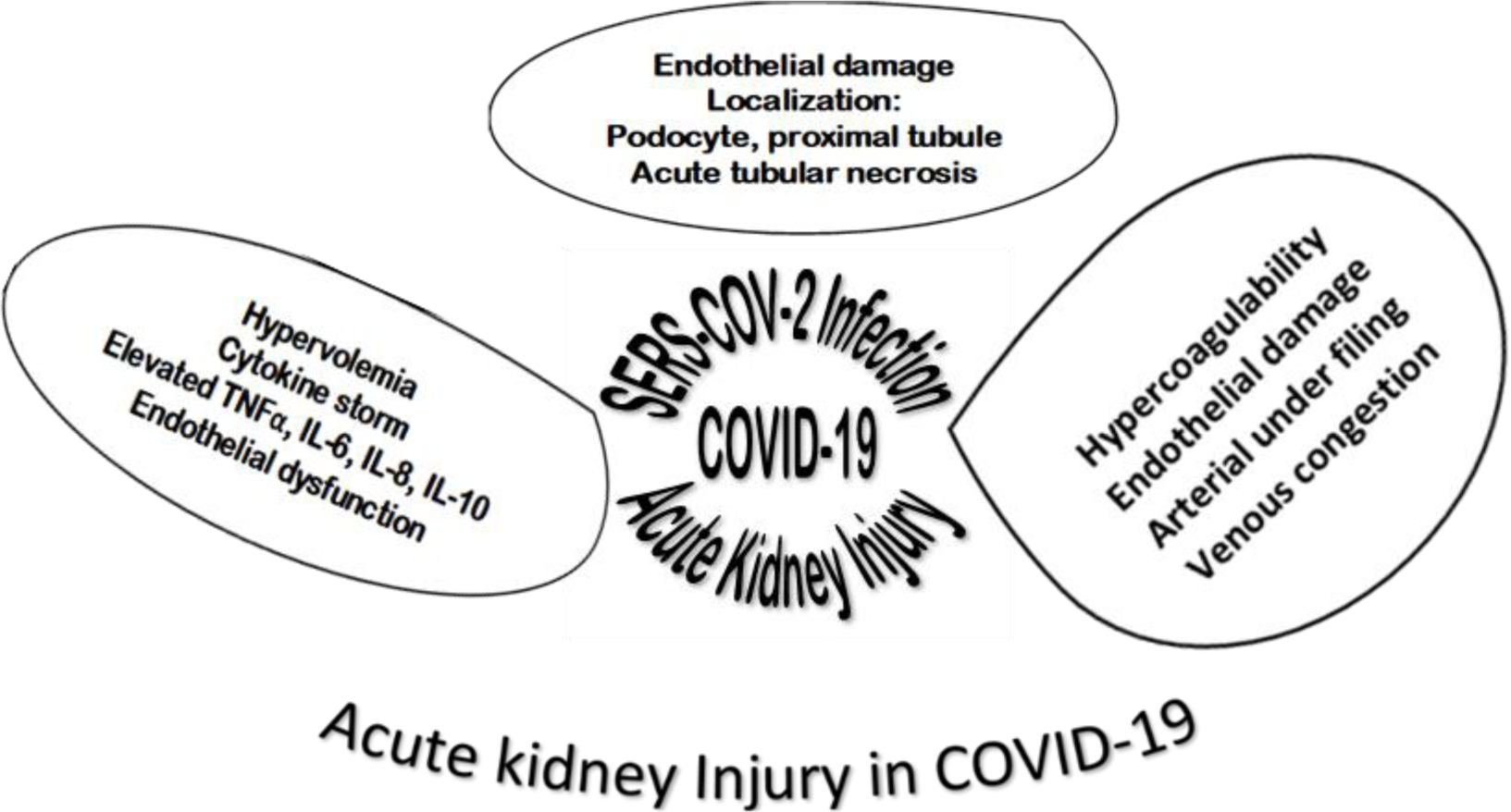
Mechanism of acute kidney injury in COVID-19 infection.

Furthermore, SARS-CoV-2 can directly infect the renal tubular epithelium and podocytes through an angiotensin converting enzyme 2 (ACE2)-dependent pathway. It causes acute tubular necrosis, mitochondrial abnormalities, the formation of protein reabsorption vacuoles, collapsing glomerulopathy, and protein leakage in Bowman’s capsule [45,46]. Another immunological mechanism is also discussed nowadays in scientific community related to AKI. Reduced lymphocyte count and elevated cytokine production is thought to be related to AKI [47,48].

## Discussions

COVID-19, the disease caused by the novel coronavirus, affected 213 countries of the world. It has infected nearly seven million people and almost half a million died. The coronavirus 2019 (COVID-19) disease outbreak occurred in Wuhan, Hubei Province, China, in December 2019, and rapidly spread to other areas of the worldwide. Now, it is well known that the novel coronavirus is dangerous than a simple respiratory virus. Usually it targets the lungs, but it can attack the whole body. Physician observed that it can cause blood clot and multi organ failure. It was observed that a certain portion of COVID-19 affected patient develop acute kidney injury with normal creatinine level. Thus, it bypasses the screening criteria and by this time the fatality occurred[6].

Our meta-analysis provided evidence that the incidence of AKI in hospitalized patients with COVID-19 infection is low, only about 8%. On the other hand, this AKI incidence during this viral infection provides a strong danger signal: the in-hospital mortality in AKI with COVID-19 infected patients is bigger, which is up to 45%. Also the fatality risk with AKI in COVID-19 infected patients is extremely high. Based on our pooled calculation it is about 4 times higher than those without AKI. Thus, clinicians should pay special attention to the treatment of COVID-19 patients with severe AKI.

We have analysed five studies and all of them reported older age and male sex were risk factors for COVID-19. This finding is consistent with other studies. In a case series studies at Wuhan Union Hospital, Beijing, China from January 29, 2020 to February 15, 2020 with COVID-19 infected 43 patients, observed older patients (≥65 years old), were more likely to have a severe type of COVID-19. Males develop more severe cases than females. In the comparable public data set of COVID-19, they also found that the percentage of older age (≥65 years) was much higher in the dead patients than who survived (83.8% in 37 deceased patients vs. 13.2% in 1,019 patients who survived) [26]. The number of men is about two and half times higher than that of women in the dead patients. Although men and women had the same vulnerability, male were more prone to dying. Therefore, gender has a vital role in case fatality with COVID-19. This gender factor, as well as higher incidences in men for most of the diseases could correlate with a general demographic fact of a shorter life expectancy in men compared to women in China and in the world in general. Other studies from Beijing Municipal Medical Taskforce informed that SARS-CoV-2 attack cells via the angiotensin-converting enzyme 2 (ACE2) receptor. It was reported earlier that high protein expression of ACE2 receptor in specific organs correlated with specific organ failures, indicated by corresponding clinical parameters in SARS patients [27, 28]. Also, it was reported earlier that circulating ACE2 levels are higher in men than in women [29]. In-hospital mortality was higher in our analyzed AKI patients. Multiple organ dysfunction including the liver, gastrointestinal tract and kidney have been reported in patients with COVID-19[24, 30]. One possible cause is that some of the COVID-19 patients had a history of chronic kidney disease (CKD). Also, it was observed by some studies that the median time period between the first symptoms and signs of COVID-19 and hospital admission was around a week [8]. By this time the patients developed a pro-inflammatory condition with abnormalities in cellular populations specially the immune cells [30]. Also, there is a higher risk for upper respiratory tract infection [31] and pneumonia. [32].Cheng et al., also explained that many COVID-19 patients could not be admitted in the early stage of disease due to lack of hospital beds. Earlier admission to hospital can help physician for management of the patient and disease spread to prevent nosocomial infection [8].

There are several evidence that severe COVID-19 patients have high level of inflammatory cytokines such as IL-1RA, IL-7, IL-8, IL-9, IL-10 [33,20]. These cytokines may contribute to develop AKI in COVID19 patients. These cytokines interact with kidney cells and trigger endothelial and tubular abnormalities. It was reported that TNF-α can bind to tubular cell receptors. As a result, it also triggers the death receptor pathway of apoptosis [34]. Also other studies experienced an elevated IL-6 in seriously ill patients with COVID-19. They observed an increased IL-6 levels in deceased patient than to survivors [35]. The lethal role of IL-6 has been demonstrated in different models [36,37]. Therefore, we can assume that these cytokines may participate to AKI in COVID-19 patients.

We found that COVID-19 patients also generally exhibited kidney disorder with elevated levels of serum creatinine, BUN followed by proteinuria & hematuria. The occurrence of kidney disorder in COVID-19 patients might be explained because of the following reasons. The SARS-CoV-2 uses angiotensin-converting enzyme 2(ACE2) as a cell entry receptor [38]. This receptor expressed in a huge amount in the small intestine, testis, kidneys, heart, thyroid, and adipose tissue [39]. Also, this receptor expressed with a much-elevated level in kidney than that in the lung [8]. Therefore, it is likely that SARS-CoV-2 can also attack kidney epithelial cells in addition to attacking lung epithelial cells [40]. This finding was supported by a recent histology report of renal tissues from autopsies and found acute renal tubular damage in six COVID-19 cases [9]. (ii) kidney disorder with other organ can enhance the inflammation process in which injury and death of renal tubular epithelial cells occurred [41,42].

### Limitation

This study has some limitations. The number of patients with AKI was small. Of the included studies a large majority are from China and the findings of this analysis may not be applicable to other part of the world. Clinical data of patient after release from hospital was absent, so we could not evaluate COVID-19 effects on long-term impact on health. More studies are needed to better investigate the risk of AKI and mortality in COVID-19 patients.

### Conclusion

Based on the available limited published data, severe AKI in patients with COVID-19 is a threatening clinical predictor and is associated with high mortality. Further studies are needed to understand the factors associated with worse outcomes among COVID-19 patients with AKI. Understanding those factors may guide care providers in making more informed dialysis eligibility decisions under conditions where resources are extremely limited.

## Abbreviations

AKI: acute kidney injury; COVID-19: Corona Virus Disease 2019; PRISMA: Preferred Reporting Items for Systematic Reviews and Meta-Analyses; MODS: Multiple Organ Dysfunction Syndromes; ARDS: Acute Respiratory Distress Syndrome; BUN: Blood Urea Nitrogen; eGFR: Estimated Glomerular Filtration Rate.

## Declarations

### Ethics approval and consent to participate

Not applicable

## Consent for publication

Not applicable.

## Availability of data and materials

Not applicable.

## Authors’ contributions

Conceptualization: MSS and RJ. Data curation: RJ. Investigation: MSS and RJ. Supervision and visualization: MSS. Writing – original draft preparation: MSS and RJ. Writing – Review & Editing: MSS and RJ. All authors have read and approved the final manuscript.

## Data Availability

All data produced in the present work are contained in the manuscript.

## Acknowledgments

Not applicable

## Authors’ information

Not applicable.

## References

1. Worldometers. COVID-19 Coronavirus Pandemic. Available from: https://www.worldometers.info/coronavirus/ [Accessed 8 June 2023].

2. Moher D, Liberati A, Tetzla J, Altman DG 02009) Preferred reporting items for systematic reviews and meta-analyses: the PRISMA statement. BMJ 339:b2535.

3. MedCalc Software. MedCalc statistical software version 16.4. 3.[computer software].

4. Lim JH, Park SH, Jeon Y, Cho JH, Jung HY, Choi JY, Kim CD, Lee YH, Seo H, Lee J, Kwon KT. Fatal Outcomes of COVID-19 in Patients with Severe Acute Kidney Injury. Journal of Clinical Medicine. 2020 Jun;9(6):1718.

5. Shao L, Li X, Zhou Y, Yu Y, Liu Y, Liu M, Zhang R, Zhang H, Wang X, Zhou F. Novel Insights Into Illness Progression and Risk Profiles for Mortality in Non-survivors of COVID-19. Frontiers in Medicine. 2020 May 22; 7:246.

6. Cheng Y, Luo R, Wang K, Zhang M, Wang Z, Dong L, Li J, Yao Y, Ge S, Xu G. Kidney disease is associated with in-hospital death of patients with COVID-19. Kidney international. 2020 Mar 20.

7. Wang D, Yin Y, Hu C, Liu X, Zhang X, Zhou S, Jian M, Xu H, Prowle J, Hu B, Li Y. Clinical course and outcome of 107 patients infected with the novel coronavirus, SARS-CoV-2, discharged from two hospitals in Wuhan, China. Critical Care. 2020 Dec; 24:1-9.

8. Li Z, Wu M, Yao J, Guo J, Liao X, Song S, Li J, Duan G, Zhou Y, Wu X, Zhou Z. Caution on kidney dysfunctions of COVID-19 patients.

9. Diao B, Feng Z, Wang C, Wang H, Liu L, Wang C, Wang R, Liu Y, Liu Y, Wang G, Yuan Z. Human kidney is a target for novel severe acute respiratory syndrome coronavirus 2 (SARS-CoV-2) infection. MedRxiv. 2020 Jan 1.

10. Li Q, Guan X, Wu P, Wang X, Zhou L, Tong Y, Ren R, Leung KS, Lau EH, Wong JY, Xing X. Early transmission dynamics in Wuhan, China, of novel coronavirus–infected pneumonia. New England Journal of Medicine. 2020 Jan 29.

11. Chan JF, Yuan S, Kok KH, To KK, Chu H, Yang J, Xing F, Liu J, Yip CC, Poon RW, Tsoi HW. A familial cluster of pneumonia associated with the 2019 novel coronavirus indicating person-to-person transmission: a study of a family cluster. The Lancet. 2020 Feb 15;395(10223):514–23.

12. Yang Y, Lu Q, Liu M, Wang Y, Zhang A, Jalali N, Dean N, Longini I, Halloran ME, Xu B, Zhang X. Epidemiological and clinical features of the 2019 novel coronavirus outbreak in China. MedRxiv. 2020 Jan 1.

13. Shimada M. BMJ Best practice, Assessment of elevated creatinine. Available from: https://bestpractice.bmj.com/topics/en-gb/935 [Accessed 13 June 2020].

14. Pagana KD, Pagana TJ. Mosby’s Manual of Diagnostic and Laboratory Tests-E-Book. Elsevier Health Sciences; 2017 Oct 8.

15. American Association for Clinical Chemistry (AACC), Estimated Glomerular Filtration Rate (eGFR). Available from: https://labtestsonline.org/tests/estimated-glomerular-filtration-rate-egfr). [Accessed 13 June 2020].

16. Huang MJ, Wei RB, Su TY, Wang Y, Li QP, Yang X, Lv XM, Chen XM. Impact of acute kidney injury on coagulation in adult minimal change nephropathy. Medicine. 2016 Nov;95(46).

17. Khatri M. Proteinuria. WebMD LLC, [Online]. Available: https://www.webmd.com/a-to-z-guides/qa/what-is-proteinuria. [Accessed 13 June 2020]. 2015.

18. Lerma EV. Proteinuria. WebMD LLC, [Online]. Available: http://emedicine.medscape.com/article/238158-overview.[Accessed 29 August 2016]. 2015.

19. Bignall ONR, 2nd, Dixon BP. Management of Hematuria in Children. Curr Treat Options Pediatr. 2018;4(3):333–49.

20. Channappanavar R, Perlman S. Pathogenic human coronavirus infections: causes and consequences of cytokine storm and immunopathology. Semin Immunopathol. 2017;39(5):529–39.

21. Conti P, Ronconi G, Caraffa AL, Gallenga CE, Ross R, Frydas I, Kritas SK. Induction of pro-inflammatory cytokines (IL-1 and IL-6) and lung inflammation by Coronavirus-19 (COVI-19 or SARS-CoV-2): anti-inflammatory strategies. J Biol Regul Homeost Agents. 2020 Mar 14;34(2):1.

22. Huang C, Wang Y, Li X, Ren L, Zhao J, Hu Y, Zhang L, Fan G, Xu J, Gu X, Cheng Z. Clinical features of patients infected with 2019 novel coronavirus in Wuhan, China. The lancet. 2020 Feb 15;395(10223):497–506.

23. Yang Y, Shen C, Li J, Yuan J, Yang M, Wang F, Li G, Li Y, Xing L, Peng L, Wei J. Exuberant elevation of IP-10, MCP-3 and IL-1ra during SARS-CoV-2 infection is associated with disease severity and fatal outcome. MedRxiv. 2020 Jan 1.

24. Chen N, Zhou M, Dong X, Qu J, Gong F, Han Y, Qiu Y, Wang J, Liu Y, Wei Y, Yu T. Epidemiological and clinical characteristics of 99 cases of 2019 novel coronavirus pneumonia in Wuhan, China: a descriptive study. The Lancet. 2020 Feb 15;395(10223):507-13.

25. Ruan Q, Yang K, Wang W, Jiang L, Song J. Clinical predictors of mortality due to COVID-19 based on an analysis of data of 150 patients from Wuhan, China. Intensive care medicine. 2020 May;46(5):846–8.

26. Jin JM, Bai P, He W, Wu F, Liu XF, Han DM, Liu S, Yang JK. Gender differences in patients with COVID-19: Focus on severity and mortality. Frontiers in Public Health. 2020 Apr 29;8:152.

27. Yang JK, Feng Y, Yuan MY, Yuan SY, Fu HJ, Wu BY, et al. Plasma glucose levels and diabetes are independent predictors for mortality and morbidity in patients with SARS. Diabet Med. 2006;23(6):623–8.

28. Yang J-K, Lin S-S, Ji X-J, Guo L-M. Binding of SARS coronavirus to its receptor damages islets and causes acute diabetes. Acta Diabetol. 2010;47(3):193–9.

29. Patel SK, Velkoska E, Burrell LM. Emerging markers in cardiovascular disease: where does angiotensin-converting enzyme 2 fit in? Clin Exp Pharmacol Physiol. 2013;40(8):551–9.

30. Betjes MG. Immune cell dysfunction and inflammation in end-stage renal disease. Nature Reviews Nephrology. 2013 May;9(5):255.

31. Cohen-Hagai K, Rozenberg I, Korzets ZE, Zitman-Gal T, Einbinder Y, Benchetrit S. Upper respiratory tract infection among Dialysis patients. Isr. Med. Assoc. J. 2016 Sep 1;18:557–60.

32. Sibbel S, Sato R, Hunt A, Turenne W, Brunelli SM. The clinical and economic burden of pneumonia in patients enrolled in Medicare receiving dialysis: a retrospective, observational cohort study. BMC nephrology. 2016 Dec 1;17(1):199.

33. Gabarre P, Dumas G, Dupont T, Darmon M, Azoulay E, Zafrani L. Acute kidney injury in critically ill patients with COVID-19. Intensive Care Medicine. 2020 Jun 12:1–0.

34. Cunningham PN, Dyanov HM, Park P, Wang J, Newell KA, Quigg RJ. Acute renal failure in endotoxemia is caused by TNF acting directly on TNF receptor-1 in kidney. The Journal of Immunology. 2002 Jun 1;168(11):5817–23.

35. Zhou F, Yu T, Du R, Fan G, Liu Y, Liu Z, Xiang J, Wang Y, Song B, Gu X, Guan L. Clinical course and risk factors for mortality of adult inpatients with COVID-19 in Wuhan, China: a retrospective cohort study. The lancet. 2020 Mar 11.

36. Su H, Lei CT, Zhang C. Interleukin-6 signaling pathway and its role in kidney disease: an update. Frontiers in immunology. 2017 Apr 21; 8:405.

37. Nechemia-Arbely Y, Barkan D, Pizov G, Shriki A, Rose-John S, Galun E, Axelrod JH. IL-6/IL-6R axis plays a critical role in acute kidney injury. Journal of the American Society of Nephrology. 2008 Jun 1;19(6):1106–15.

38. Zhou P, Yang XL, Wang XG, Hu B, Zhang L, Zhang W, Si HR, Zhu Y, Li B, Huang CL, Chen HD. A pneumonia outbreak associated with a new coronavirus of probable bat origin. nature. 2020 Mar;579(7798):270-3.

39. Li MY, Li L, Zhang Y, Wang XS. Expression of the SARS-CoV-2 cell receptor gene ACE2 in a wide variety of human tissues. Infectious diseases of poverty. 2020 Dec;9:1–7.

40. Yeung ML, Yao Y, Jia L, Chan JF, Chan KH, Cheung KF, Chen H, Poon VK, Tsang AK, To KK, Yiu MK. MERS coronavirus induces apoptosis in kidney and lung by upregulating Smad7 and FGF2. Nature microbiology. 2016 Feb 22;1(3):1–8.

41. Faubel S, Edelstein CL. Mechanisms and mediators of lung injury after acute kidney injury. Nature Reviews Nephrology. 2016 Jan;12(1):48.

42. Wang H, Ma S. The cytokine storm and factors determining the sequence and severity of organ dysfunction in multiple organ dysfunction syndrome. The American journal of emergency medicine. 2008 Jul 1;26(6):711–5.

43. Ronco C, Reis T, Husain-Syed F. Management of acute kidney injury in patients with COVID-19. The Lancet Respiratory Medicine. 2020 May 14.

44. Varga Z, Flammer AJ, Steiger P, Haberecker M, Andermatt R, Zinkernagel AS, Mehra MR, Schuepbach RA, Ruschitzka F, Moch H. Endothelial cell infection and endotheliitis in COVID-19. The Lancet. 2020 May 2;395(10234):1417–8.

45. Su H, Yang M, Wan C, Yi LX, Tang F, Zhu HY, Yi F, Yang HC, Fogo AB, Nie X, Zhang C. Renal histopathological analysis of 26 postmortem findings of patients with COVID-19 in China. Kidney international. 2020 Apr 9.

46. Larsen CP, Bourne TD, Wilson JD, Saqqa O, Moh’d A S. Collapsing Glomerulopathy in a Patient With COVID-19. Kidney international reports. 2020 Jun 1;5(6):935–9.

47. Zhou F, Yu T, Du R, Fan G, Liu Y, Liu Z, Xiang J, Wang Y, Song B, Gu X, Guan L. Clinical course and risk factors for mortality of adult inpatients with COVID-19 in Wuhan, China: a retrospective cohort study. The lancet. 2020 Mar 11.

48. Ronco C, Reis T. Kidney involvement in COVID-19 and rationale for extracorporeal therapies. Nature Reviews Nephrology. 2020 Apr 9:1–3.

